# Rapid nanopore metagenomic sequencing and predictive susceptibility testing of positive blood cultures from intensive care patients with sepsis

**DOI:** 10.1101/2023.06.15.23291261

**Authors:** Patrick N. A. Harris, Michelle J. Bauer, Lukas Lüftinger, Stephan Beisken, Brian M. Forde, Ross Balch, Menino Cotta, Luregn Schlapbach, Sainath Raman, Kiran Shekar, Peter Kruger, Jeff Lipman, Seweryn Bialasiewicz, Lachlan Coin, Jason A. Roberts, David L. Paterson, Adam D. Irwin

## Abstract

**Background:** Direct metagenomic sequencing from positive blood culture (BC) broths, to identify bacteria and predict antimicrobial susceptibility, has been previously demonstrated using Illumina-based methods, but is relatively slow. We aimed to evaluate this approach using nanopore sequencing to provide more rapid results.

**Methods:** Patients with suspected sepsis in 4 intensive care units were prospectively enrolled. Human-depleted DNA was extracted from positive BC broths and sequenced using nanopore (MinION). Species abundance was estimated using Kraken2, and a cloud-based artificial intelligence (AI) system (AREScloud) provided *in silico* antimicrobial susceptibility testing (AST) from assembled contigs. These results were compared to conventional identification and phenotypic AST.

**Results:** Genus-level agreement between conventional methods and metagenomic whole genome sequencing (MG-WGS) was 96.2% (50/52), but increased to 100% in monomicrobial infections. In total, 262 high quality AREScloud AST predictions across 24 samples were made, exhibiting categorical agreement (CA) of 89.3%, with major error (MA) and very major error (VME) rates of 10.5% and 12.1%, respectively. Over 90% CA was achieved for some taxa (e.g. *Staphylococcus aureus*), but was suboptimal for *Pseudomonas aeruginosa* (CA 50%). In 470 AST predictions across 42 samples, with both high quality and exploratory-only predictions, overall CA, ME and VME rates were 87.7%, 8.3% and 28.4%. VME rates were inflated by false susceptibility calls in a small number of species / antibiotic combinations with few representative resistant isolates. Time to reporting from MG-WGS could be achieved within 8-16 hours from blood culture positivity.

**Conclusions:** Direct metagenomic sequencing from positive BC broths is feasible and can provide accurate predictive AST for some species and antibiotics, but is sub-optimal for a subset of common pathogens, with unacceptably high VME rates. Nanopore-based approaches may be faster but improvements in accuracy are required before it can be considered for clinical use. New developments in nanopore sequencing technology, and training of AI algorithms on larger and more diverse datasets may improve performance.

## INTRODUCTION

Sepsis is a major cause of morbidity and mortality. Rapid pathogen identification and antimicrobial susceptibility phenotyping is critical to selection of appropriate treatment and ensuring optimal patient outcomes (1, 2). Current pathogen identification and culture-based antimicrobial susceptibility testing (AST) can take up to 3 days, or longer. Consequently, rapid molecular detection and gene profiling methodologies are needed (2–4), especially in an era of an increasing prevalence of antimicrobial resistance.

We have previously demonstrated the application of Illumina-based sequencing from positive blood culture broths (5). This approach showed reasonable performance in both species-level identification and predictive AST from metagenomic data, but there were few advantages over conventional methods in terms of turn-around times to clinical reporting. In this study, we aimed to evaluate the use of nanopore sequencing, using a similar approach and an established DNA extraction method, to determine whether reductions in turn-around times can be achieved without sacrificing diagnostic performance. We compared a machine-learning based whole genome sequencing predictive AST tool to conventional culture-based methods, in order to determine whether such methods could be applicable in a diagnostic laboratory and achieve acceptable performance characteristics.

## METHODS

This was a sub-study of the DIRECT program: a prospective, observational multicentre study of children and adults presenting to the intensive care unit (ICU) with clinical features of sepsis (6). Patients were screened for enrolment in four ICUs (3 adult, 1 paediatric) in Brisbane, Australia (Royal Brisbane and Women’s Hospital, The Prince Charles Hospital, Princess Alexandra Hospital and Queensland Children’s Hospital). Patients who met inclusion criteria and none of the exclusion criteria were eligible for enrolment (Table 1).

**Table 1:** Inclusion and exclusion criteria

| ***Inclusion criteria*** | ***Exclusion criteria*** |
| --- | --- |
| a) Age >1 month | a) inability to gain informed consent |
| b) admitted to ICU | b) neonates (<1 month age) |
| c) decision to treat for suspected sepsis† | c) imminent death likely |
| d) blood cultures collected within 12h | d) palliative care intent |
| e) commenced on IV antibiotics within 24h, or a change in antibiotics initiated within 24h for a new episode of infection | e) on renal replacement therapy * |
|  | f) use of extra-corporeal membrane oxygenation (ECMO) * |

\* necessary as the main study included therapeutic drug monitoring and dose optimisation
† defined as suspected / proven infection with evidence of end organ dysfunction

### Ethics

Ethics approval was granted by the Children’s Health Queensland Hospital and Health Service Human Research Ethics Committee (HREC) [HREC/19/QCHQ/55177], with governance approvals for all participating sites. Written informed consent was obtained from all participants (or their parent or legal guardian). Approval was granted by the Queensland Civil and Administrative Tribunal [CRL024-19] to include patients unable to consent for themselves under the Guardianship and Administration Act 2000.

### Sampling

This study included patients with positive blood cultures and presence of bacteria confirmed by Gram stain and microscopy. Samples with non-bacterial species (e.g. *Candida*) were excluded from further analysis (as the analysis pipelines are optimised for prokaryotic organisms). Samples with likely contamination (e.g. mixed coagulase-staphylococci), which were not worked-up further by the clinical lab for identification or susceptibility testing, were also not analysed further. For some samples, susceptibility testing was not routinely performed on cultured isolates (e.g. anaerobes), hence predictive AST was not assessed.

Blood culture bottles (FA plus, FN plus and paediatric PF plus bottles; bioMérieux) were removed from BACT/Alert Virtuo System once flagged positive with microbial growth on Gram stain. A de-identified 10 mL aliquot of the positive blood culture broth was processed by a blinded researcher in a separate research lab (The University of Queensland, Centre for Clinical Research), located on the same campus, within 1.5 hours (Figure 1). An uninoculated blood culture broth was also sampled to determine the extent of background DNA contamination. Methods for DNA extraction have been detailed previously (5). In brief, host genomic DNA (gDNA) was depleted using the MolYsis Complete and MolYsis Basic kits (Molzym, Germany) 0.2mL and 1mL protocols, respectively, according to manufacturer’s instructions, (with minor modifications), then centrifuged at 10,000g for 30 seconds and the supernatant removed (5). The microbial pellet then underwent further gDNA extraction using UltraClean kits. Samples were extracted for genomic DNA upon receipt and the remaining sample frozen at −80°C, and if required, thawed to room temperature from frozen.

**Figure 1:**
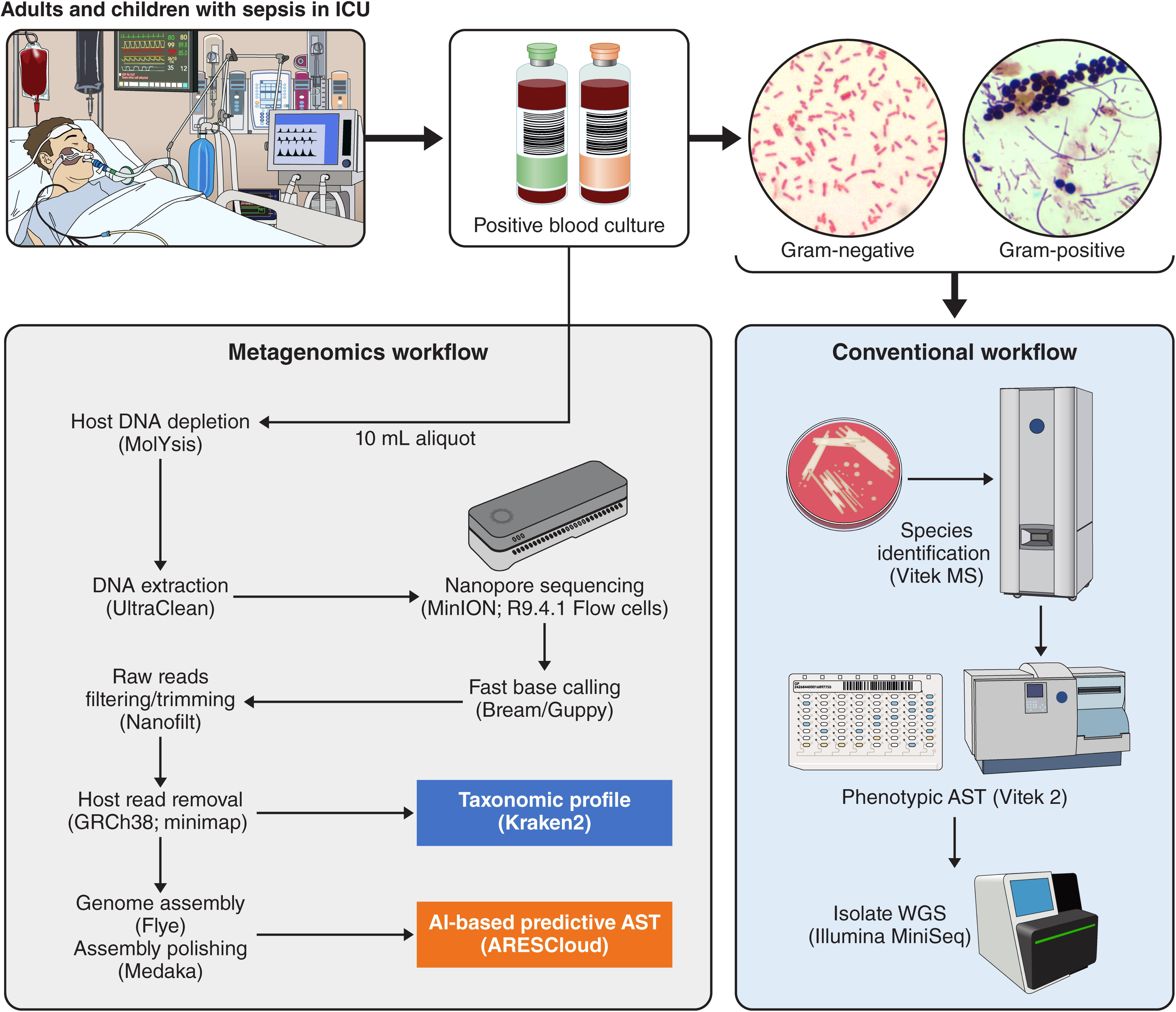
Workflow for metagenomic and conventional analysis of positive blood cultures

DNA quality and purity checks were undertaken using the QUBIT fluorometer (Life Technologies), NanoDrop 2000 Spectrophotometer (Thermo Scientific) and Agilent TapeStation 4150 using Genomic DNA ScreenTape and Reagents. For comparison, cultured isolates from the positive BC broths were also retrieved from the clinical laboratory for WGS (Figure 1).

### Sequencing

Libraries were prepared using the Rapid Barcoding Sequencing or Nanopore Genomic DNA Ligation kit with Native barcoding according to manufacturer’s instructions (Oxford Nanopore Technologies). Libraries were loaded into the R9.4.1 flow cell and run on the MinION MK1B device. After flow cell quality checks, all sequencing utilised the flow cell priming kit (EXP-FLP002) and sequencing commenced at −180mV; voltage drift was accounted for where the flow cell went through a wash protocol. The sequencing runs were monitored with MinKNOW version 21.05.25 core 4.3.12. Fast basecalling model was applied with Bream version 6.2.6 and Guppy version 5.0.16 (version 3.2.9 used prior to February 2021). Libraries were run for 72 hours. Final run QC analysis was undertaken with PycoQC prior to bioinformatic analysis. In addition, pure colonies of bacteria isolated from positive blood cultures were also sequenced, using DNA extracted by QIAGEN DNeasy Ultraclean Microbial Kit, with quantification by Life Technologies QUBIT fluorometer and library preparation with Illumina ILMN DNA LP tagmentation. Library size was determined by Agilent TapeStation 4150 using D1000 High Sensitivity kit. Sequencing was performed on the Illumina MiniSeq platform with the 300 cycle output reagent kit (Figure 1).

### Metagenome assemblies, taxonomic profiling and WGS-AST using machine-learning

Assembly and binning for WGS-AST from metagenomes was performed using a previously published workflow (7). Raw reads were trimmed and quality filtered using Nanofilt 2.8 (8), and mapped against the GRCh38 genome using minimap 2.24 (9) to remove host reads. Where possible given input sequencing depth, retained reads were assembled with flye 2.9 (10) and parameters “--nano-raw --meta -- iterations 3”. Whole metagenome assemblies were polished with Oxford Nanopore Medaka 1.6.1 and parameters “-m r941_min_fast_g303” (11, 12). Binning of assembled metagenomes into metagenomic bins was performed with MaxBin 2.2.7 (13) and MetaBAT 2.15 (12). Bins were unified using DASTool 1.1.5 (14). Resulting bins were post-processed to improve retainment of AMR marker genes from high quality unbinned contigs, as previously described (7). Taxonomy was assigned using Kraken2 (15) and visualised using Krona plots (https://fordegenomics.github.io/direct). Completeness and duplication of bins was assessed with BUSCO (16) and QUAST (17). For each sample, no more than one resulting bin had genome quality metrics compatible with downstream AST prediction. Downstream analysis was thus performed on whole metagenome assemblies to reduce loss of AMR information in the binning process. Metagenome assemblies were uploaded to the AREScloud web application, release 2022-10 (Ares Genetics GmbH, Vienna, AT) for genomic prediction of antimicrobial susceptibility. The platform used stacked classification machine learning (ML) WGS-AST models trained on ARESdb (18), combined with rule-based resistance prediction via ResFinder 4 (19) to provide species-specific susceptibility/resistance (S/R) predictions. If no high-quality ML models were available in AREScloud for certain taxa, non-specific ResFinder 4 calls based only on generalized presence of antibiotic resistance genes were used, but were flagged as being lower confidence predictions. Where sequence data from cultured isolates and paired BC broth samples were available, *in silico* resistance gene profiles were determined by screening the draft assembled genomes against the NCBI resistance gene database using AMRFinderPlus (version 3.10.24) (20) with default parameters (90% sequence identity and 90% sequence coverage) and compared for concordance in the presence / absence of AMR genes across the sample types.

AST predictions for a total of 25 antibiotic compounds were generated, where appropriate and relevant for that species. True negatives (TN) were defined as data points where both the reference method (phenotypic AST) and the test method (AREScloud) returned a negative (i.e. susceptible) result; true positives (TP) where both methods returned a positive (i.e. resistant) result; false positives (FP) where the reference method returned a negative and the tested method returned a positive result; false negatives (FN), where the reference method returned a positive and the tested method returned a negative result. Very major error (VME) and major error (ME) rates were defined following CLSI M52 guidelines (21) as the fraction of cases identified as resistant by the reference method which were identified as susceptible by the tested method (FN / (FN +TP)), and the fraction of cases identified as susceptible by the reference method which were identified as resistant by the tested method (FP / (FP + TN)), respectively. Categorical agreement (CA) between results of WGS-AST and conventional AST were calculated (CA = (TN + TP) / (TN + FP + FN + TP)) for antimicrobial-organism combinations.

### Conventional species identification and AST

All genomics based species identification and AST results were compared to conventional phenotypic methods validated for clinical use at Pathology Queensland. Species identification was performed using MALDI-TOF (Vitek MS, bioMérieux) on pure cultured isolates, with AST performed by Vitek 2 automated broth microdilution (N-246 AST cards; bioMérieux), using EUCAST clinical breakpoints applicable at the time (22). For certain species (e.g. *Streptococcus pyogenes*) AST was undertaken using disk diffusion according to EUCAST methods (23), or by Etest (bioMérieux) where appropriate (e.g. penicillin for *Streptococcus pneumoniae*). For some species where EUCAST breakpoints were not available (e.g. *Aeromonas* spp.), CLSI breakpoints were applied. Conventional phenotypic testing reported for clinical use by the diagnostic laboratory was considered the reference standard against which genomic results were compared.

## RESULTS

### Blood Culture Microorganisms

A total of 66 positive blood culture samples, from 201 enrolled patients, demonstrated bacterial growth, from which 52 were included for further MG-WGS analysis, with exclusions reflecting non-bacterial growth (e.g. *Candida* sp.), missing samples or likely contaminants (e.g. mixed coagulase-negative staphylococci) that were not worked up further by the clinical laboratory (Figure 2; Supplementary Table S1). Samples included 27 gram-positive and 23 gram-negative bacterial species, with 2 samples showing polymicrobial growth (Table 2).

**Figure 2:**
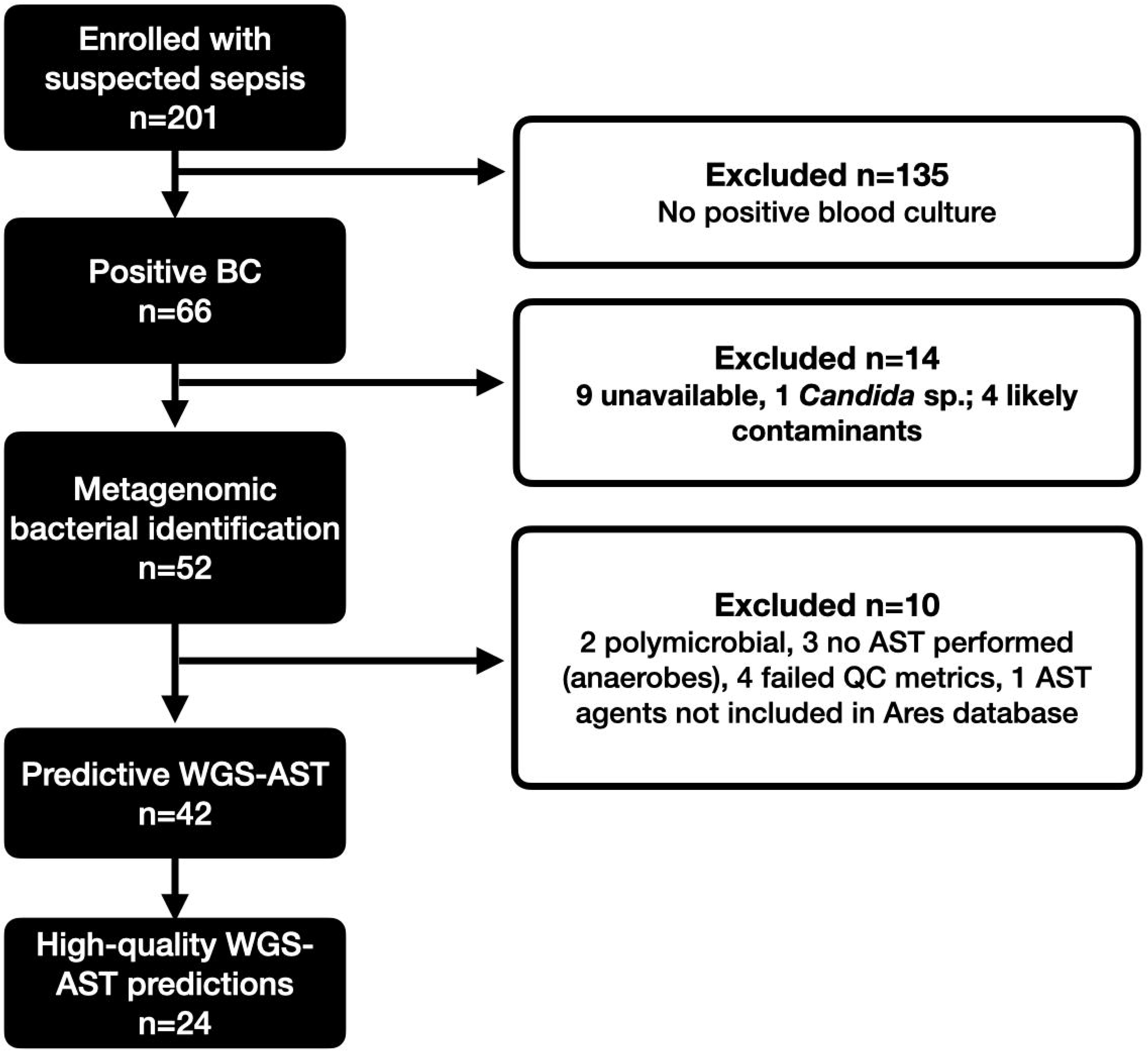
Flow diagram for sample inclusion. BC = blood culture; WGS-AST = whole genome sequencing antimicrobial susceptibility testing; QC = quality control

**Table 2:** Species identification by conventional methods

| Species | N |
| --- | --- |
| **Gram-positive (N=27)** |  |
| *Staphylococcus aureus* | 10 |
| *Staphylococcus epidermidis* | 7 |
| *Streptococcus pneumoniae* | 2 |
| *Streptococcus pyogenes* | 1 |
| Group C *Streptococcus* | 1 |
| *Staphylococcus capitis* | 1 |
| *Staphylococcus lugdunensis* | 1 |
| *Staphylococcus haemolyticus* | 1 |
| *Enterococcus faecalis* | 1 |
| *Clostridium perfringens* | 1 |
| *Eggerthella lenta* | 1 |
| **Gram-negative (N=23)** |  |
| *Escherichia coli* | 7 |
| *Enterobacter cloacae* complex | 4 |
| *Klebsiella (Enterobacter) aerogenes* | 1 |
| *Pseudomonas aeruginosa* | 3 |
| *Klebsiella pneumoniae* | 3 |
| *Haemophilus influenzae* | 1 |
| *Stenotrophomonas maltophilia* | 1 |
| *Aeromonas* sp. | 1 |
| *Prevotella intermedia* | 1 |
| *Ochrobactrum anthropi* | 1 |
| **Polymicrobial (N=2)** |  |
| *Escherichia coli* + *Enterobacter cloacae* | 1 |
| *Proteus mirabilis* + *Staphylococcus hominis* | 1 |
| **Total** | **52** |

### Taxonomy identification of metagenomic samples and *in silico* predictive AST

Samples were run either on a single flow cell, or multiplexed with up to 12 samples per flow cell, with a median sequencing yield per flow cell of 2.5 Gbp for multiplexed samples, and 1.5 Gbp for single samples. Taxonomic identification of metagenomic samples yielded excellent agreement to genus level compared to conventional methods (50/52, 96.2%); for monomicrobial samples agreement was 100% (50/50). In two samples, genus-level agreement was obtained, with sequencing provided a more refined identification; the pipeline identified species belonging to *Enterobacter cloacae* complex (*E. hormaechei* for sample 9420-58 and *E. asburiae* for sample 9420-32) in two samples reported as *E. cloacae* by phenotypic methods. The correct species level identity resulting from MG-WGS of both samples was confirmed by sequencing of the cultured isolates. For one polymicrobial sample the secondary pathogen (*E. cloacae*) reported by phenotypic methods was not apparent in the Kraken2 report of input reads, where only one of the cultured pathogens (*E. coli*) was identified. For another polymicrobial sample, the presence of *Staphylococcus hominis* was identified by phenotypic methods alongside *Proteus mirabilis*, but only ∼ 15% of bacterial reads in the sample could be matched to genus *Staphylococcus* and were insufficient for predictive MG-AST. For one sample (9421-30), no identification was achieved due to inadequate input data; with only 145 very short reads (mean read length 191), thus no further processing was possible. Sequencing from an uninoculated blood culture broth revealed a very low number of reads (n=28) mapping to bacterial genomes (e.g. *E. coli*), compared to a mean number of reads of 271411 mapping to bacterial taxa for positive culture broths included in the MG-AST analysis.

A total of 470 phenotypic AST results with matched metagenomic AST calls were analysed. As conventional AST was not routinely performed in all samples (e.g. likely contaminants, anaerobic organisms), predictive MG-AST was only compared where phenotypic results were available. In addition, 2 polymicrobial samples were excluded. For an additional 6 samples, neither exploratory nor high quality MG-AST predictions, were available (4 samples had insufficient reads for assembly and in 2 samples, antibiotics reported by phenotypic methods, were not available in the AREScloud database). As such, a total of 42 samples had both MG-AST and phenotypic AST calls compared, and for 24 samples, high quality AREScloud predictions were available.

Overall CA was 89.3% for 262 AST results across 24 samples for which quality MG-AST predictions were available, including 7 common BSI organisms (*Staphylococcus aureus, Enterococcus faecalis, Escherichia coli, Klebsiella aerogenes, Klebsiella pneumoniae, Streptococcus pneumoniae* and *Pseudomonas aeruginosa*), but with 12.1% VMEs (mainly seen in *E. coli* with tobramycin and trimethoprim-sulfamethoxazole, driven by a single *E. coli* sample with poor assembly metrics) and 10.5% MEs (mainly seen in *P. aeruginosa* against cefepime, ciprofloxacin, gentamicin and meropenem, *K. aerogenes* against ceftazidime and ceftriaxone, *E. coli* against amikacin, cephazolin, ampicillin and cefoxitin, and *K. pneumoniae* against cephazolin and ciprofloxacin) (Tables 3 and 5; Supplementary Table S2).

**Table 3:** Performance of predictive MG-AST by species compared to Vitek 2, for samples with high quality predictions

| Species | N | Categorical Agreement (%) | Major Error (%) | Very Major Error (%) |
| --- | --- | --- | --- | --- |
| *Staphylococcus aureus* | 9 | 97.2 | 3.2 | - |
| *Escherichia coli* | 6 | 87.8 | 9.2 | 23.5 |
| *Klebsiella pneumoniae* | 3 | 93.8 | 6.5 | 0 |
| *Pseudomonas aeruginosa* | 3 | 52.4 | 47.6 | - |
| *Enterococcus faecalis* | 1 | 100 | 0 | - |
| *Klebsiella aerogenes* | 1 | 83.3 | 20 | 0 |
| *Streptococcus pneumoniae* | 1 | 66.7 | 33.3 | - |
| **Overall** | **24** | **89.3** | **10.5** | **12.1** |

Note: blank cells reflect no data for calculations (e.g. no resistance seen in that species by reference method)

For all 470 AST predictions across 42 samples, including both high-quality and exploratory-only results, CA was 87.7%, with 28.4% VME and 8.3% MEs (Table 4). VMEs were mainly seen with one isolate of *Aeromonas* sp. (100%; false susceptibility for amoxicillin-clavulanate, meropenem and trimethoprim), *Pseudomonas aeruginosa* (100%; false susceptibility for ticarcillin-clavulanate), *E. coli* (31.6%; false susceptibility for tobramycin, trimethoprim-sulfamethoxazole and ticarcillin-clavulanate), *Ochrobactrum anthropi* (33.3%; false susceptibility for ceftriaxone), *Staphylococcus haemolyticus* (50%; false susceptibility for cephalothin, ciprofloxacin, rifampicin, tetracycline) and *Staphylococcus epidermidis* (20%; false susceptibility for cephalothin, ciprofloxacin and teicoplanin). MEs were seen in *Pseudomonas aeruginosa* (45.5%; false resistance for cefepime, ceftazidime, ciprofloxacin, meropenem) and *Klebsiella pneumoniae* (11.1%, false resistance for cephazolin, cefoxitin and ciprofloxacin) (Supplementary Table S2). Not all of the tested compounds achieved satisfactory performance even when high quality predictions were achieved, with CA ranging from >95% (for amoxicillin-clavulanate, gentamicin, erythromycin, fusidic acid, ticarcillin-clavulanate and vancomycin) to as low as 55.6% for cephazolin, with high rates of VMEs for trimethoprim-sulfamethoxazole (66.7%), trimethoprim (25%) and tobramycin (100%) (Table 6). The only agents that would pass acceptance criteria (>95% CA, <3% ME and <1.5% VME) even when only using high quality predictions, would be amoxicillin-clavulanate, erythromycin and fusidic acid. While vancomycin and ticarcillin-clavulanate had 100% CA and no MEs, the lack of resistant isolates precluded calculation of the rate of VMEs.

**Table 4:** Performance of predictive MG-AST by species compared to Vitek 2, for all samples including those with both high confidence and exploratory-only predictions

| **Species** | **N** | **Categorical Agreement (%)** | **Major Error (%)** | **Very Major Error (%)** |
| --- | --- | --- | --- | --- |
| *Staphylococcus aureus* | 9 | 96.6 | 2.9 | 7.7 |
| *Staphylococcus epidermidis* | 7 | 85.5 | 7.1 | 35 |
| *Escherichia coli* | 6 | 86.5 | 8.7 | 30 |
| *Enterobacter cloacae* | 3 | 92.3 | 7.4 | 8.3 |
| *Klebsiella pneumoniae* | 3 | 90 | 11.1 | 0 |
| *Pseudomonas aeruginosa* | 3 | 50 | 45.5 | 100 |
| *Aeromonas* sp. | 1 | 70 | 0 | 100 |
| *Enterococcus faecalis* | 1 | 100 | 0 | 0 |
| *Haemophilus influenzae* | 1 | 100 | 0 | - |
| *Klebsiella aerogenes* | 1 | 93.3 | 9.1 | 0 |
| *Ochrobactrum anthropi* | 1 | 87.5 | 0 | 33.3 |
| *Staphylococcus capitis* | 1 | 83.3 | 0 | 50 |
| *Staphylococcus haemolyticus* | 1 | 67.7 | 0 | 57.1 |
| *Staphylococcus lugdunensis* | 1 | 100 | 0 | 0 |
| *Streptococcus pneumoniae* | 1 | 75 | 25 | - |
| *Streptococcus pyogenes* | 1 | 100 | 0 | 0 |
| *Streptococcus dysgalactiae* | 1 | 100 | 0 | - |
| **Overall** | **42** | **87.7** | **8.3** | **28.4** |

Note: blank cells reflect no data for calculations (e.g. no resistance seen in that species by reference method)

**Table 5:** Performance of MG-AST compared to Vitek MS, by species and compound, where high quality predictions were available

| Species | Compound | CA (%) | ME (%) | VME (%) |
| --- | --- | --- | --- | --- |
| *Enterococcus faecalis* | Gentamicin | 100 | 0 | - |
|  | Erythromycin | 100 | - | 0 |
|  | Linezolid | 100 | 0 | - |
|  | Teicoplanin | 100 | 0 | - |
|  | Vancomycin | 100 | 0 | - |
| *Escherichia coli* | Amikacin | 83.3 | 16.7 | - |
|  | Amoxicillin + clavulanic acid | 100 | 0 | - |
|  | Ampicillin | 83.3 | 33.3 | 0 |
|  | Cefazolin | 40 | 75 | 0 |
|  | Cefepime | 100 | 0 | 0 |
|  | Cefoxitin | 83.3 | 16.7 | - |
|  | Ceftazidime | 100 | 0 | 0 |
|  | Ceftriaxone | 100 | 0 | 0 |
|  | Ciprofloxacin | 100 | 0 | 0 |
|  | Gentamicin | 100 | 0 | 0 |
|  | Meropenem | 100 | 0 | - |
|  | Sulfamethoxazole + trimethoprim | 60 | 0 | 66.7 |
|  | Tobramycin | 83.3 | 0 | 100 |
|  | Trimethoprim | 83.3 | 0 | 25 |
| *Klebsiella (Enterobacter) aerogenes* | Amikacin | 100 | 0 | - |
|  | Amoxicillin + clavulanic acid | 100 | - | 0 |
|  | Cefazolin | 100 | - | 0 |
|  | Ceftazidime | 0 | 100 | - |
|  | Ceftriaxone | 0 | 100 | - |
|  | Ciprofloxacin | 100 | 0 | - |
|  | Gentamicin | 100 | 0 | - |
|  | Meropenem | 100 | 0 | - |
|  | Sulfamethoxazole + trimethoprim | 100 | 0 | - |
|  | Ticarcillin + clavulanic acid | 100 | 0 | - |
|  | Tobramycin | 100 | 0 | - |
|  | Trimethoprim | 100 | 0 | - |
| *Klebsiella pneumoniae* | Amikacin | 100 | 0 | - |
|  | Amoxicillin + clavulanic acid | 100 | 0 | 0 |
|  | Cefazolin | 66.7 | 33.3 | - |
|  | Ceftazidime | 100 | 0 | - |
|  | Ceftriaxone | 100 | 0 | - |
|  | Ciprofloxacin | 50 | 50 | - |
|  | Gentamicin | 100 | 0 | - |
|  | Meropenem | 100 | 0 | - |
|  | Sulfamethoxazole<br>+trimethoprim | 100 | 0 | - |
|  | Ticarcillin + clavulanic acid | 100 | 0 | - |
|  | Tobramycin | 100 | 0 | - |
|  | Trimethoprim | 100 | 0 | - |
| *Pseudomonas aeruginosa* | Amikacin | 100 | 0 | - |
|  | Cefepime | 33.3 | 66.7 | - |
|  | Ceftazidime | 0 | 100 | - |
|  | Ciprofloxacin | 33.3 | 66.7 | - |
|  | Gentamicin | 66.7 | 33.3 | - |
|  | Meropenem | 33.3 | 66.7 | - |
|  | Tobramycin | 100 | 0 | - |
| *Staphylococcus aureus* | Benzylpenicillin | 100 | 0 | 0 |
|  | Ciprofloxacin | 100 | 0 | - |
|  | Clindamycin | 88.9 | 14.3 | 0 |
|  | Erythromycin | 100 | 0 | 0 |
|  | Fusidic acid | 100 | 0 | - |
|  | Gentamicin | 100 | 0 | - |
|  | Linezolid | 100 | 0 | - |
|  | Mupirocin | 88.9 | 11.1 | - |
|  | Rifampicin | 88.9 | 11.1 | - |
|  | Teicoplanin | 100 | 0 | - |
|  | Tetracycline | 100 | 0 | - |
|  | Vancomycin | 100 | 0 | - |
| *Streptococcus pneumoniae* | Benzylpenicillin | 0 | 100 | - |
|  | Erythromycin | 100 | 0 | - |
|  | Vancomycin | 100 | 0 | - |
| **Overall** | **All agents** | **89.3** | **10.5** | **12.1** |

Note: blank cells reflect no data for calculations (e.g. no susceptibility / resistance seen in that species by reference method)

**Table 6:** Performance of MG-AST compared to Vitek MS for all compounds, where high quality predictions were available

| Compound | CA (%) | ME (%) | VME (%) |
| --- | --- | --- | --- |
| Amikacin | 92.3 | 7.7 | - |
| Amoxicillin + clavulanic acid | 100 | 0 | 0 |
| Ampicillin | 83.3 | 33.3 | 0 |
| Benzylpenicillin | 90 | 50 | 0 |
| Cefazolin | 55.6 | 57.1 | 0 |
| Cefepime | 77.8 | 25 | 0 |
| Cefoxitin | 83.3 | 16.7 | - |
| Ceftazidime | 69.2 | 33.3 | 0 |
| Ceftriaxone | 88.9 | 12.5 | 0 |
| Ciprofloxacin | 85.7 | 15 | 0 |
| Clindamycin | 88.9 | 14.3 | 0 |
| Erythromycin | 100 | 0 | 0 |
| Fusidic acid | 100 | 0 | - |
| Gentamicin | 95.5 | 4.8 | 0 |
| Linezolid | 100 | 0 | - |
| Meropenem | 83.3 | 16.7 | - |
| Sulfamethoxazole + trimethoprim | 77.8 | 0 | 66.7 |
| Ticarcillin + clavulanic acid | 100 | 0 | - |
| Tobramycin | 92.3 | 0 | 100 |
| Trimethoprim | 90 | 0 | 25 |
| Vancomycin | 100 | 0 | - |
| **Overall** | **89.3** | **10.5** | **12.1** |

Note: blank cells reflect no data for calculations (e.g. no resistance seen in that species by reference method)

Detection of AMR genes from nanopore generated assemblies of bacterial genomes from BC broths showed some discrepancies compared to Illumina-based sequencing of pure cultured isolates (Figure 3), although several of these reflected more specific allele calls from sequencing pure isolates (e.g. *bla*_CTX-M_ from MG-WGS, but *bla*_CTX-M-15_ from pure isolate). Illumina-based WGS of pure isolates detected a median of 3 additional AMR gene targets (range -1 to 14; IQR 1-5). In only a single sample did nanopore detect one more AMR gene target than Illumina.

**Figure 3:**
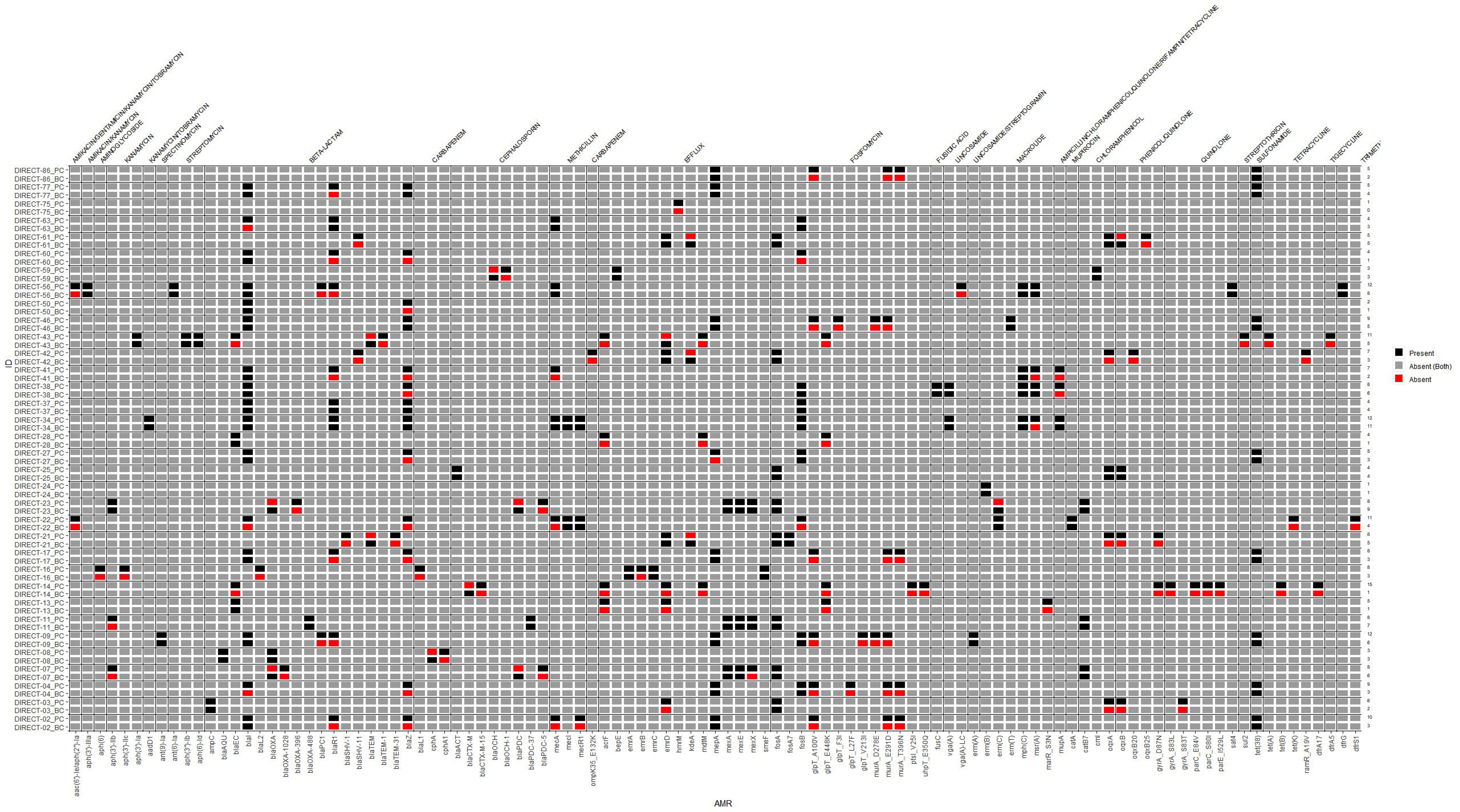
Heat map comparing antimicrobial resistance genes detected from Nanopore-generated sequences from blood culture broth extractions, compared to Illumina-generated sequences from pure cultured isolates from the same sample

In terms of potential turn-around times for direct nanopore sequencing from blood cultures; pre-sequencing steps took ∼4 hours, including: a) host DNA reduction, ∼2 hours, b) DNA Extraction, ∼30 minutes, c) DNA amplification, ∼1 hour, and d) DNA Library prep, ∼30 minutes. Adequate data for downstream processing would usually be achieved within ∼4 hours, but flow cells were run up to 12 hours for single samples, and 72 hours for multiplexed samples. The predictive MG-AST calls from AREScloud were available within ∼ 1 hour. As such, the potential turn-around time from flagging of a positive blood culture to report generation, could be achieved within 9-17 hours, potentially faster than conventional AST methods and Illumina-based methods (up to 48 hours, or longer).

## DISCUSSION

We describe a nanopore-based MG-WGS approach using positive blood culture broth for pathogen detection and taxonomic classification In monomicrobial infections, the performance is encouraging, with 100% agreement to genus level. Polymicrobial samples remain challenging, with only one of two pathogens identified in samples encountered in this study. In two samples with species belonging to the *E. cloacae* complex, MG-WGS identification was more accurate than conventional methods, although discrimination by MALDI-TOF of these species is known to be problematic, without additional analysis (24, 25). Sequencing directly from blood samples to detect pathogenic bacteria in patients with sepsis and bloodstream infection, is limited by low loads of bacterial DNA in blood at the time of presentation, high concentrations of human DNA and challenges in discriminating background low-level contaminating DNA (26, 27). Adding a culture-amplification step by sequencing from positive blood culture broths, as described in this study, increases the amount of bacteria DNA available for sequencing.

A key advance in the application of metagenomic diagnostics direct from clinical samples, would be the ability to accurately predict antimicrobial susceptibility, independently of conventional culture-based methods. However, the presence or absence of resistance genes does not always predict the phenotype, which may be modified by gene expression, gene copy number and other post-translational effects (28). The machine-learning algorithm used in this study is based on a large sample bank with matched whole genome sequenced clinical isolates and AST results collected from several international centres (18). Such an approach has advantages in that the algorithm does not require a clear understanding of the association between genotype and phenotype, but will learn to use relevant genomic features if supplied with adequate amounts of data.

The use of direct metagenomic sequencing and WGS-AST from positive blood culture broths holds some promise, and nanopore long-read sequencing using MinION offers potential time advantages over Illumina-based approaches. Accuracy of pathogen identification was similar to results previously achieved with Illumina (95% species-level agreement) (5). However, using nanopore sequencing chemistries, flow cells and base-callers available at the time of study, MG-AST predictions based on nanopore data were considerably less accurate than Illumina-based methods, where CA of >95% was demonstrated for common gram-negative pathogens against 17 antimicrobials (with an overall 11% VME rate) (5). Nanopore sequencing in some samples resulted in significant fragmentation and incompleteness of assembled genomes, caused by an insufficient number of reads and low average read length in a subset of data, as well as lower depth of sequencing compared with Illumina. In addition, reads overall exhibited low per-base accuracy (average Phred score of 10.48, i.e. approximately 90% accuracy), likely due to base-calling with the “fast” profile and the use of an older versions of the Guppy base-caller (v5.0.16; and v3.2.9 used in earlier sequencing runs). Additional work is needed to assess the performance of current nanopore consumables (e.g. R10.4.1 flow cells and kit 14 chemistry), which are reported to achieve very high sequencing accuracies (29), and more current base-calling software. The high rates of MEs/VMEs encountered in common species in this study, would currently preclude application for clinical use. CLSI M52 guidelines recommend that new AST systems demonstrate CA ≥90% and rates of MEs and VMEs <3% (21), although given the high risk of VMEs to patient care, the FDA stipulates VME rates to be <1.5% (30). However, it should be noted that some of these errors occurred in less critical or uncommonly prescribed species/antibiotic combinations (e.g. *E. coli* and tobramycin). It is hoped that errors could also be mitigated by database enhancement and training of the algorithms on a larger number and broader range of organisms. The application of sequencing from blood cultures may also allow faster identification of slow-growing or fastidious organisms. One potential approach to reduce time to reporting, might be the early sampling of blood culture broths, before they flag positive on automated detections systems. In this way, there may be adequate pathogen load to undertake sequencing, while reducing the overall turn-around time.

Limitations to this study are acknowledged. While samples included in the study were prospectively collected, and included most common species causing sepsis, a more extensive range of pathogens, including diverse AMR phenotypes would need to be assessed to understand the reliability, broader applicability and clinical utility of this approach. Furthermore, our collection included few samples with resistance to some agents, inflating VME rate in some species/antibiotic combinations. For example, there were only two *Pseudomonas aeruginosa* isolates with resistance to any antimicrobial tested; with resistance to ticarcillin-clavulanate that were both falsely reported as susceptible by MG-AST, leading to a 100% VME rate (albeit only from low-quality exploratory predictions). The absence of any resistances in the *Pseudomonas* samples to other agents precluded the ability to calculate any VMEs for other antibiotics using high-quality predictions. It is also acknowledged that Vitek 2 AST is not a reference method for MIC determination (such as broth microdilution or agar dilution) and is an imperfect standard for comparison, despite being commonly used in clinical diagnostic laboratories. Additionally, improvements in nanopore flow cell technology have occurred since this study was performed, including the possibility of adaptive sequencing that can actively exclude human DNA during the sequencing process (31), which may also further improve the application of these methods.

## Conclusions

Direct metagenomic sequencing from positive blood culture broths in patients with sepsis is feasible and can provide accurate species-level identification of causative pathogens, especially in monomicrobial infections. Predictive AST shows promise for some bacterial species and antibiotic combinations, but is sub-optimal for a number of common pathogens with unacceptably high VME rates driven by less reliable calls for certain species and drug combinations. Diagnostic performance characteristics were marginally improved by only accepting results where high-quality predictions were available. Nanopore-based approaches may be faster and provide data in real time, but improvements in accuracy across a broader range of organisms are required before it can be considered for clinical use. Improved performance should be achievable with training of ML algorithms on larger and more diverse datasets, and masking of results where poor performance of certain species and drug combinations are recognised. Furthermore, ongoing developments in the accuracy of rapid sequencing technologies, should lead to improved performance of these methods for eventual diagnostic use.

## Data Availability

.

## Acknowledgements and funding

This work was funded by a grant from the Queensland Genomics Health Alliance (QGHA; subsequently Queensland Genomics) clinical implementation, innovation and incubation program and by a Brisbane Diamantina Health Partners Health System Improvement Ideas Grant (MRFF Rapid Applied Research Translation Program). Patrick Harris was supported by an Early Career Fellowship from the National Health and Medical Research Council (GNT1157530). Jason Roberts would like to acknowledge funding from the Australian National Health and Medical Research Council for a Centre of Research Excellence (APP2007007) and an Investigator Grant (APP2009736) as well as an Advancing Queensland Clinical Fellowship. Adam Irwin is supported by a National Health and Medical Research Council Investigator grant (GNT1197743). We would like to acknowledge the input and support of Thom Cuddihy, Cameron Buckley, Jason Meyer, Kara Brady, Cheryl Fourie, Natalie Sharp, Luminita Vlad, Scott A. Beatson, Julia Clark, Krispin Hajkowicz, Haakon Bergh and David Whiley.

## Data availability

Raw sequence reads have been uploaded to NCBI under Bioproject PRJNA982891. Taxonomic classifications for blood culture samples can be visualised here: https://fordegenomics.github.io/direct

